# Longitudinal Evaluation of Harlem United’s Multiservice Model on Clinical, Behavioral, and Social Outcomes Among Clients Living with HIV

**DOI:** 10.64898/2026.05.23.26353941

**Authors:** Briana S Monk, Danielle Strauss

**Affiliations:** Department of Epidemiology and Biostatistics, CUNY Graduate School of Public Health; Department of Epidemiology and Biostatistics, Data Analytics & Program, Harlem United

**Keywords:** Longitudinal, Generalized estimating equation (GEE), PHQ4, Integrated model, Social determinants of health (SDOH)

## Abstract

**Background/Objectives:** People living with HIV face overlapping hardship through medical, behavioral, and social needs that require an integrated and coordinated approach. Harlem United’s multiservice model provides healthcare, food assistance, housing support, harm reduction services, behavioral health counseling, case management, and much more to support their clients. This study is an examination on how the participation in Harlem United’s multiservice model is associated with changes over time in client health, behavioral health, and social outcomes.

**Methods:** This study performed a longitudinal program evaluation examining Harlem United clients enrolled between January 2020 and January 2025 who remained engaged in services for a minimum of one year. Client outcomes were assessed across three time points: Baseline, Year 1, and Year 2. The sample included 154 clients at baseline (N=154) with a total of 428 observations (N=428). Quantitative measures that were assessed included program involvement, housing stability, PHQ4 scores, food insecurity, medication adherence, and viral suppression. Data was analyzed using IBM SPSS Statistics through descriptive statistics, frequency tables, and generalized estimating equation models (GEE) to account for repeated observation over time.

**Results:** Medication adherence and viral suppression remained consistently high across all time points in the longitudinal study suggesting that most clients were virally suppressed or undetectable at baseline. Housing stability was statistically significant Wald X2 (2) = 156.073, p < 0.001 with improvements noted in Year 1 and Year 2 compared to baseline. Program level was significantly associated with PHQ4 scores, Wald X2 (1) = 7.902, p = 0.005. Food insecurity was also associated with PHQ4 scores, Wald X2 (1) = 5.462, p = 0.019. Findings suggest that clients with higher PHQ4 scores were involved in more programs compared to clients only enrolled in 1-2 programs. Additionally, clients with higher PHQ4 scores were more food insecure highlighting the relationship between social needs and mental health.

**Conclusion:** Findings suggest that Harleam United’s multiservice model played a supportive role in the maintenance of health and social outcomes through medication adherence and viral suppression. Although, significant improvement was not reflected across several outcomes, the association between PHQ4 scores, food insecurity, and an increase in program involvement suggest that the multiservice is reaching more clients with complex behavioral and social needs. Continued integration of these services is important for sustaining client stability while addressing social determinants of health.

## Introduction

Achieving health equity among HIV patients requires focusing on societal efforts to address inequalities as well as historical and contemporary injustices. Social determinants of health and disparities continue to threaten public health outcomes among populations disproportionately affected by HIV.^1^ “Approximately 1.2 million people in the U.S. are living with HIV” yet a significant number remain unaware of their status increasing risk of transmission.^3^ Minority communities are more likely to face higher rates of HIV infection and poorer health outcomes based on conditions where they live, work, and have access to healthcare.^2^ Factors that can limit access to quality health care are associated through stigma, discrimination, and unstable housing resulting in reduced engagement in HIV prevention and treatment services, impacting viral suppression and long-term health.^1^ In 2022, “Black/African American persons made up approximately 12% of the population of the United States but accounted for 37% (11,900) of the estimated 31,800 new HIV infections.”^3^ “White persons made up 61% of the population of the United States but accounted for 24% (7,600) of new HIV infections.”^3^ “Hispanic/Latino persons made up 18% of the population of the United States but accounted for 33% (10,500) of HIV infections.^3^ “Together Black/African Americans and Hispanic/Latino people made up more than half (70%) of estimated new HIV infections in 2022.”^3^

In addition to disparities in incidence, only 65% of people living with HIV in the U.S. will achieve viral suppression status.^3^ Many of the new HIV transmissions are linked to individuals who are either undiagnosed or not engaged in HIV care further emphasizing the role of supportive services and engagement in reducing HIV transmission.^3^ Research has emphasized that a more comprehensive approach is needed to address HIV disparities.^2^ A population-level approach that targets the drivers of inequity can improve outcomes in marginalized communities reinforcing the need for both clinical and social needs to be met.^2^

Harlem United operates an integrated multiservice model designed to address the intersecting needs of individuals living with HIV including those experiencing homelessness, behavioral health challenges, and substance use.^4^ This organization delivers care across four domains: healthcare, housing, harm reduction, and supportive services with the goal of addressing social determinants of health and improving health outcomes for individuals living with HIV.^4^ At the clinical level, Harlem United provides comprehensive health services including primary care, dental care, psychiatry, and specialty services.^16^ Behavioral health services are delivered through individual and group psychotherapy, crisis intervention, and medication management with specialized support offered for LGBTQIA individuals.^16^ These services can be offered both virtually and in person to ensure accessibility and continuity of care.^16^

One of the most used services within the multiservice model is the permanent and temporary housing.^16^ It serves as an intervention to provide stability for clients experiencing chronic homelessness and HIV.^16^ Harm reduction is employed through Harlem United to recognize how substance use plays a factor in influencing HIV outcomes.^16^ Services include syringe exchange programs, overdose prevention training, medication-assisted treatment, as well as access to HIV and STI testing and treatment.^16^ In addition to clinical, housing, and harm reduction, Harlem United delivers supportive services through case management and care coordination.^16^ Harlem United’s multiservice model outlines how a holistic, person-centered framework can combine medical care with social and structural needs. This model aims to improve viral suppression, reduce PHQ4 scores, and enhance the overall quality of life among vulnerable populations living with HIV.

Since the use of antiretroviral therapy, HIV care has significantly advanced leading to longer life.^7–9^ However, for many patients living with HIV, additional factors such as mental health and housing instability coexist, further influencing long-term health.^7–9^ Clinical care alone is not enough to sustain HIV outcomes which is why integrated care models are needed to address the full range of clinical, behavioral and social needs.^7–10^

Integrated care is essential for people living with HIV because the relationship between mental health and HIV can be complex.^7–10^ Systematic reviews have provided evidence that integrated HIV care and mental health have the potential to improve both patient outcomes and service delivery, yet different delivery models provide different outcomes.^9,14,17,18^ It is still difficult to determine which delivery model is most effective and for whom.^9,14,17,18^ Existing research acknowledges the value that integrated care brings, yet continues to only examine specific components of care leaving behind social outcomes.^12–14,17,18^ Coordinated strategies that address all relevant services such as harm reduction, housing support, and food assistance are needed though community based HIV programs to allow patients to be more engaged and supported within their services.^11–14^ Additionally, neighborhood-level approaches such as community centers highlight why integration is necessary.^11–14^ This approach for patients living with HIV is more conducive to their surroundings as an individual to see how their health is influenced by their environment.^11–14^ Despite the growing evidence, there is still insufficient longitudinal data on the impact of integrated HIV programs. Continued research is needed to demonstrate the benefit of these models in either improvements or maintenance of health and social outcomes. The objective of this longitudinal study is to address this gap by evaluating Harlem United’s multiservice tool and examining outcomes across time periods from Baseline, Year 1, and Year 2. This evaluation seeks to examine how integrated services impact people living with HIV.

## Methods

The study design used quantitative data aimed at performing a longitudinal analysis through a generalized estimating equations (GEE) model. This model was selected to evaluate change over time and assess the effectiveness of Harlem United’s multiservice model. The GEE model was the most appropriate model because it accounts for repeated measures within clients and captured trends to better understand how program engagement, or number of programs, influence clinical, behavioral, and social outcomes. Data received from Harlem United was de-identified with all data being reported through an eICare number to protect the client’s confidentiality and adhere to all data privacy and confidentiality standards. Per the human protection program at CUNY Graduate School of Public Health & Health Policy, IRB approval is not needed.

Data was collected through Harlem United’s internal program database that collected data across multiple platforms through surveys. Data was merged into an excel document to be further cleaned and analyzed. The data was classified into four categories: client outcomes, behavioral health, social determinants of health, and program engagement. Client outcomes were assessed through viral suppression and medication adherence. Behavioral health was further identified through PHQ4 scores (psychological distress) and TIC scores (trauma-related symptoms). Social determinants of health were noted through food insecurity, housing status, and history of homelessness. Program engagement referred to the number of programs a client was involved with throughout the duration of the program. Lastly, demographics demonstrated the covariates present in the study through race, gender, ethnicity, and sexual orientation.

Study participants were selected as clients that have been enrolled in services at Harlem United beginning in January 2020 or later and have been enrolled for a minimum of one year to compare to the baseline assessment. Longitudinal data was assessed from three time points: Baseline, Year 1, and Year 2. Any time points after Year 2 were excluded due to small sample size. The total sample size of Harlem United clients was N=428 with baseline clients N=154.

The statistical software used to conduct the data analysis is IBM SPSS Statistics. Descriptive statistics were used to identify demographic variables. Chi-square tests were used to examine positive or negative associations between variables such viral load and medication adherence, mental health and food insecurity, and TIC and PHQ4 scores. Different levels of distribution were used in the GEE model based on data being binary, ordinal, or nominal. Binomial logit GEE models were best used for binary variables such as viral suppression, stable housing, and medication adherence. Poisson GEE was best used for count outcomes such as PHQ4 scores and TIC screener scores. Each model evaluated for time, time x program level, or program level to determine the effects of the multiservice model.

## Results

Table 1 presents baseline demographic characteristics from the Harlem United Study sample. The sample included 154 clients at baseline. Of the 154 clients, 104 clients were Black/African American representing 67.5% of the total sample with white clients accounting for 16.2%. Clients identifying as Other Race represented 12.3% with smaller proportions noted among clients who identified as Indian/Alaska Native, Asian, Hawaiian/Pacific Islander, more than one race or did not know their race category.

**Table 1:** Harlem United Baseline Demographics.

|  |  | Count | Column N % |
| --- | --- | --- | --- |
| Race1 | American Indian/Alaska Native | 2 | 1.3% |
|  | Asian | 1 | 0.6% |
|  | Black/African American | 104 | 67.5% |
|  | Don't Know | 1 | 0.6% |
|  | Hawaiian/Pacific Islander | 1 | 0.6% |
|  | More than one Race | 1 | 0.6% |
|  | Other Race | 19 | 12.3% |
|  | White | 25 | 16.2% |
| Gender | Female | 40 | 26.0% |
|  | Male | 106 | 68.8% |
|  | Other/Non-Conforming | 1 | 0.6% |
|  | Transgender ID - Female | 7 | 4.5% |
| Sexual Orientation | Bisexual | 5 | 3.2% |
|  | Choose not to Respond | 4 | 2.6% |
|  | Gay | 32 | 20.8% |
|  | Heterosexual | 101 | 65.6% |
|  | Other/Non-Conforming | 4 | 2.6% |
|  | Pansexual | 1 | 0.6% |
|  | Something else | 1 | 0.6% |
|  | Unknown | 6 | 3.9% |
| Ethnicity1 | Don't Know | 6 | 3.9% |
|  | Hispanic/Latino | 35 | 22.7% |
|  | Non-Hispanic/Latino | 113 | 73.4% |

**Table 2:**
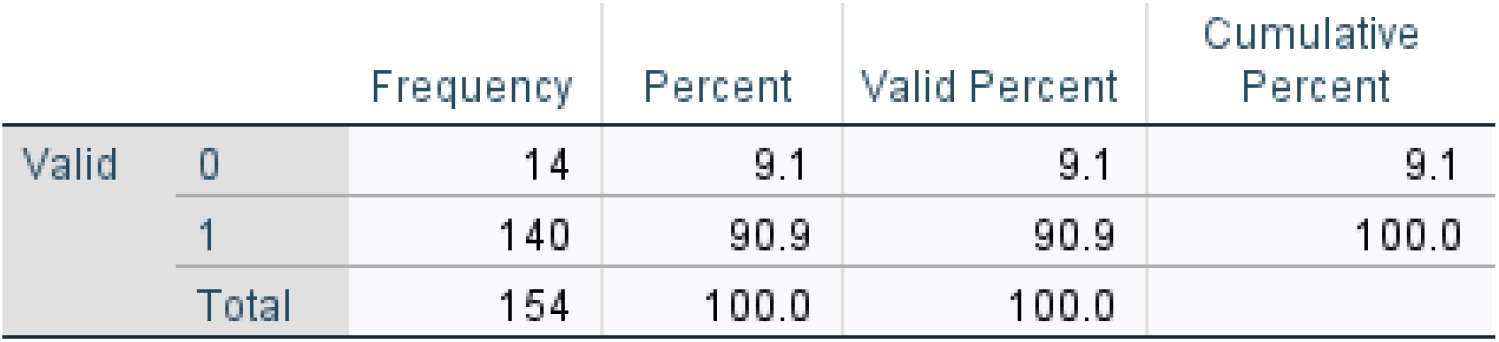
Homelessness at Baseline.

**Table 3:**
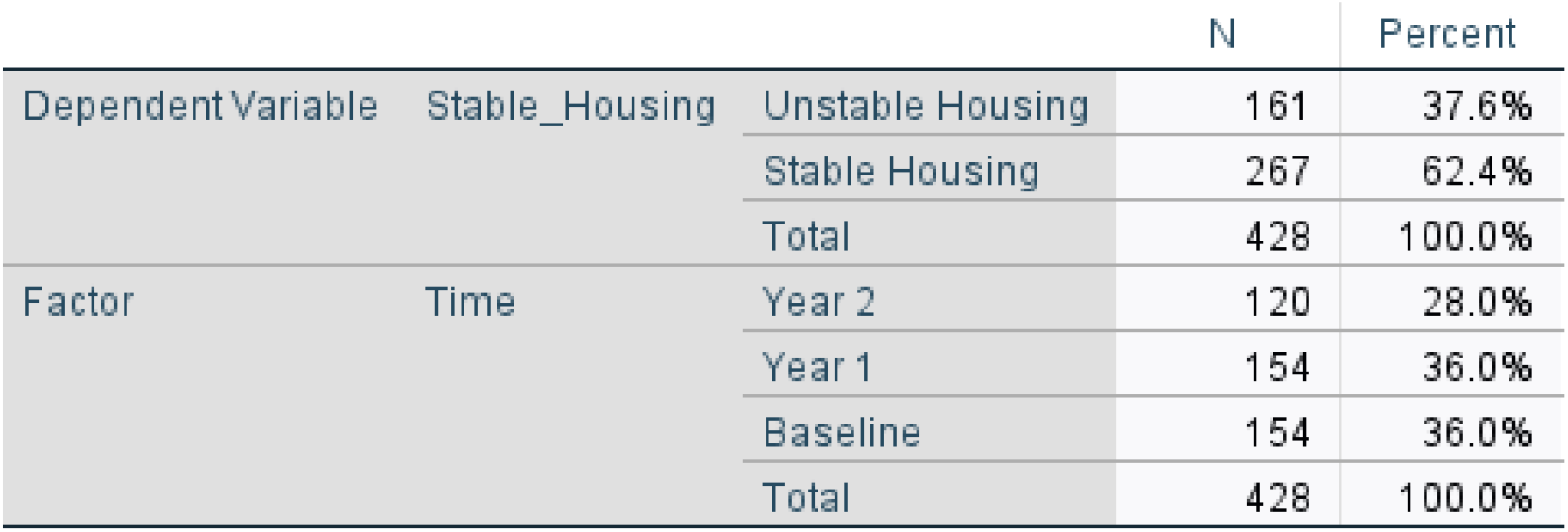
Categorical Variable Information.

**Table 3a:** Tests of Model Effects.

| Source | Type III |  |  |
| --- | --- | --- | --- |
|  | Wald Chi-Square | df | Sig. |
| (Intercept) | 16.075 | 1 | <.001 |
| Time | 156.073 | 2 | <.001 |

Dependent Variable: Stable\_Housing
Model: (Intercept), Time

**Table 3b:** Parameter Estimates.

| Parameter | B | Std. Error | 95% Wald Confidence Interval |  | Hypothesis Test |  |  | Exp(B) | 95% Wald Confidence Interval for Exp(B) |  |
| --- | --- | --- | --- | --- | --- | --- | --- | --- | --- | --- |
|  |  |  | Lower | Upper | Wald Chi-Square | df | Sig. |  | Lower | Upper |
| (Intercept) | -2.155 | .2641 | -2.672 | -1.637 | 66.564 | 1 | <.001 | .116 | .069 | .195 |
| [Time=3] | 4.553 | .4014 | 3.766 | 5.339 | 128.665 | 1 | <.001 | 94.875 | 43.204 | 208.345 |
| [Time=2] | 4.538 | .3714 | 3.811 | 5.266 | 149.332 | 1 | <.001 | 93.548 | 45.176 | 193.716 |
| [Time=1] | 0<sup>a</sup> | . | . | . | . | . | . | 1 | . | . |
| (Scale) | 1 |  |  |  |  |  |  |  |  |  |

Dependent Variable: Stable\_Housing
Model: (Intercept), Time
a. Set to zero because this parameter is redundant.

**Table 4:** Medication Aherence Categorical Variable Information.

|  |  |  | N | Percent |
| --- | --- | --- | --- | --- |
| Dependent Variable | Adherence_Binary | 0 | 39 | 17.5% |
|  |  | 1 | 184 | 82.5% |
|  |  | Total | 223 | 100.0% |
| Factor | Time | Year 2 | 73 | 32.7% |
|  |  | Year 1 | 79 | 35.4% |
|  |  | Baseline | 71 | 31.8% |
|  |  | Total | 223 | 100.0% |
|  | Program_Level | 3-5 Programs | 57 | 25.6% |
|  |  | 1-2 Programs | 166 | 74.4% |
|  |  | Total | 223 | 100.0% |

**Table 4a:**
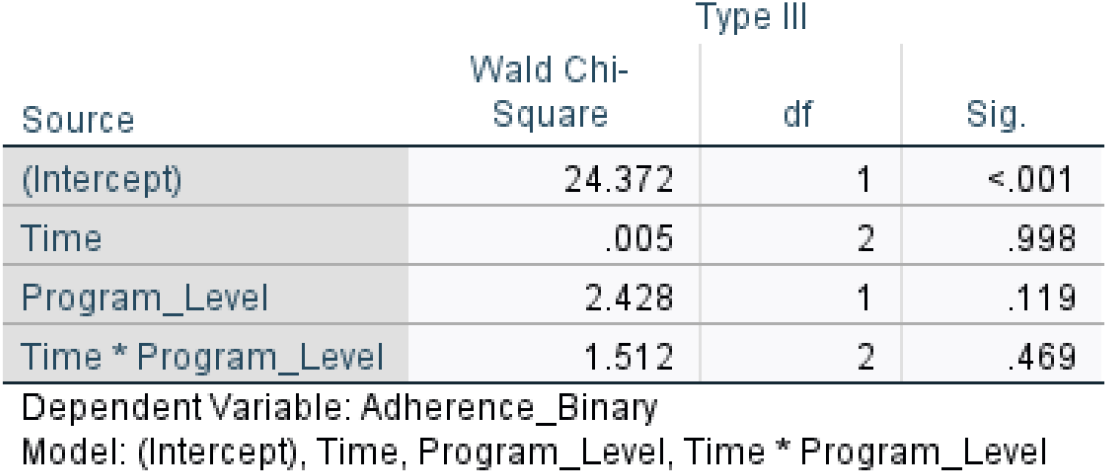
Medication Adherence Tests of Model Effects.

| Source | Wald Chi-Square | Type III |  |
| --- | --- | --- | --- |
|  |  | df | Sig. |
| (Intercept) | 24.372 | 1 | <.001 |
| Time | .005 | 2 | .998 |
| Program_Level | 2.428 | 1 | .119 |
| Time * Program_Level | 1.512 | 2 | .469 |

Dependent Variable: Adherence\_Binary
Model: (Intercept), Time, Program\_Level, Time \* Program\_Level

**Table 4b:** Medication Adherence Parameter Estimates.

| Parameter | B | Std. Error | 95% Wald Confidence Interval |  | Hypothesis Test |  |  | Exp(B) | 95% Wald Confidence Interval for Exp(B) |  |
| --- | --- | --- | --- | --- | --- | --- | --- | --- | --- | --- |
|  |  |  | Lower | Upper | Wald Chi-Square | df | Sig. |  | Lower | Upper |
| (Intercept) | 1.792 | .3819 | 1.043 | 2.540 | 22.014 | 1 | <.001 | 6.000 | 2.839 | 12.683 |
| [Time=3] | .223 | .3764 | -.515 | .961 | .351 | 1 | .553 | 1.250 | .598 | 2.614 |
| [Time=2] | -.077 | .2301 | -.528 | .374 | .112 | 1 | .738 | .926 | .590 | 1.454 |
| [Time=1] | 0<sup>a</sup> | . | . | . | . | . | . | 1 | . | . |
| [Program_Level=2.00] | -.780 | .6977 | -2.148 | .587 | 1.250 | 1 | .263 | .458 | .117 | 1.799 |
| [Program_Level=1.00] | 0<sup>a</sup> | . | . | . | . | . | . | 1 | . | . |
| [Time=3] * [Program_Level=2.00] | -.473 | .6542 | -1.755 | .810 | .522 | 1 | .470 | .623 | .173 | 2.247 |
| [Time=3] * [Program_Level=1.00] | 0<sup>a</sup> | . | . | . | . | . | . | 1 | . | . |
| [Time=2] * [Program_Level=2.00] | .164 | .5104 | -.836 | 1.164 | .103 | 1 | .748 | 1.178 | .433 | 3.204 |
| [Time=2] * [Program_Level=1.00] | 0<sup>a</sup> | . | . | . | . | . | . | 1 | . | . |
| [Time=1] * [Program_Level=2.00] | 0<sup>a</sup> | . | . | . | . | . | . | 1 | . | . |
| [Time=1] * [Program_Level=1.00] | 0<sup>a</sup> | . | . | . | . | . | . | 1 | . | . |
| (Scale) | 1 |  |  |  |  |  |  |  |  |  |

Dependent Variable: Adherence\_Binary
Model: (Intercept), Time, Program\_Level, Time \* Program\_Level
a. Set to zero because this parameter is redundant.

**Table 5:** PHQ4 Scores & Food Insecurity Categorical Variable Information.

|  |  |  | N | Percent |
| --- | --- | --- | --- | --- |
| Factor | Time | Year 2 | 113 | 28.0% |
|  |  | Year 1 | 149 | 36.9% |
|  |  | Baseline | 142 | 35.1% |
|  |  | Total | 404 | 100.0% |
|  | Program_Level | 3-5 Programs | 101 | 25.0% |
|  |  | 1-2 Programs | 303 | 75.0% |
|  |  | Total | 404 | 100.0% |
|  | Food_Insecurity | Food Insecure | 63 | 15.6% |
|  |  | Food Secure | 341 | 84.4% |
|  |  | Total | 404 | 100.0% |

**Table 5a:** Tests of Model Effects.

| Source | Wald Chi-Square | Type III |  |
| --- | --- | --- | --- |
|  |  | df | Sig. |
| (Intercept) | 21.893 | 1 | <.001 |
| Time | 4.342 | 2 | .114 |
| Program_Level | 7.902 | 1 | .005 |
| Time * Program_Level | 4.930 | 2 | .085 |
| Food_Insecurity | 5.462 | 1 | .019 |

Dependent Variable: Q13\_PHQ-4 Score
Model: (Intercept), Time, Program\_Level, Time \* Program\_Level, Food\_Insecurity

**Table 5b:**
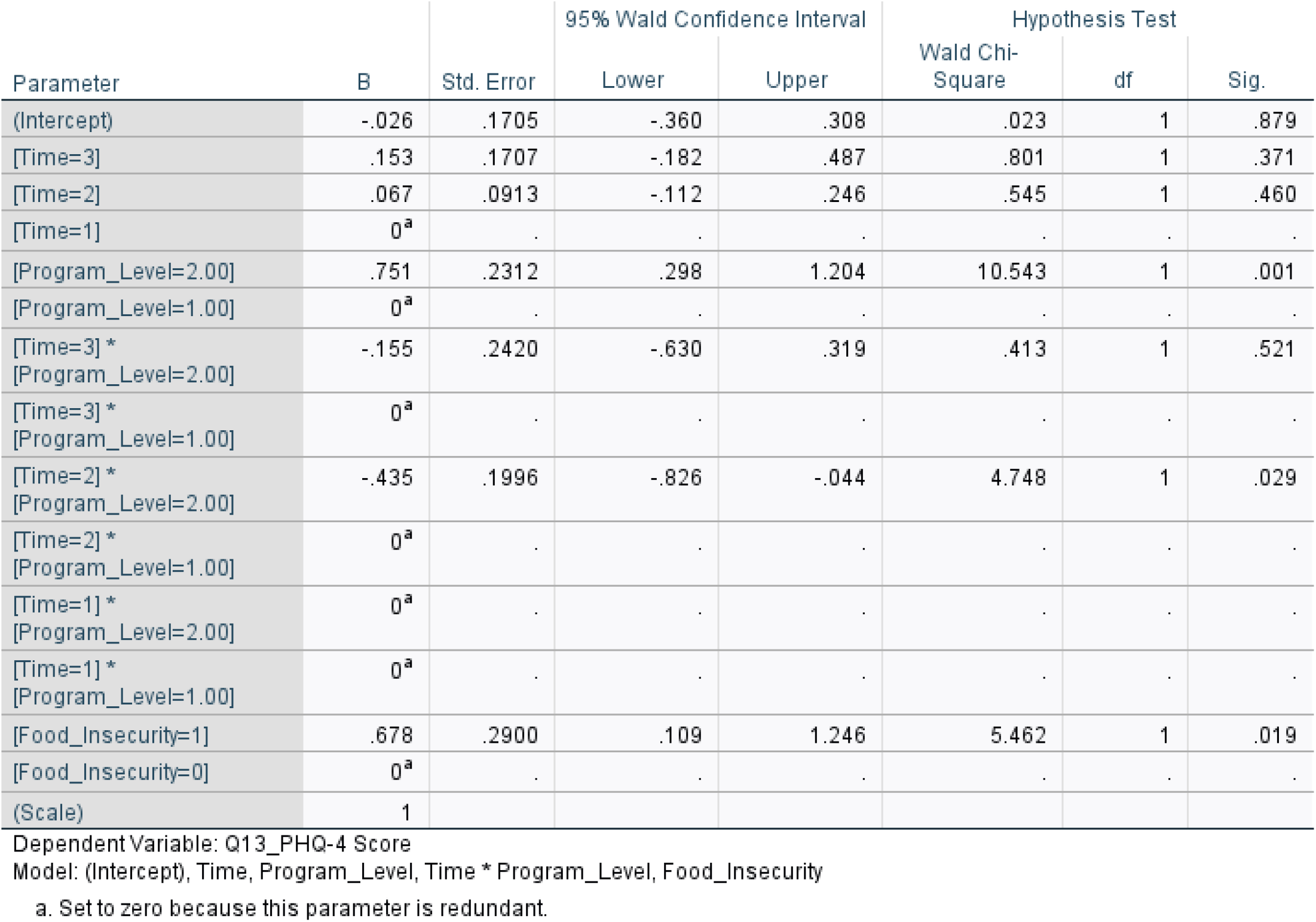
Parameter Estimates.

| Parameter | B | Std. Error | 95% Wald Confidence Interval |  | Hypothesis Test |  |  |
| --- | --- | --- | --- | --- | --- | --- | --- |
|  |  |  | Lower | Upper | Wald Chi-Square | df | Sig. |
| (Intercept) | -.026 | .1705 | -.360 | .308 | .023 | 1 | .879 |
| [Time=3] | .153 | .1707 | -.182 | .487 | .801 | 1 | .371 |
| [Time=2] | .067 | .0913 | -.112 | .246 | .545 | 1 | .460 |
| [Time=1] | 0<sup>a</sup> | . | . | . | . | . | . |
| [Program_Level=2.00] | .751 | .2312 | .298 | 1.204 | 10.543 | 1 | .001 |
| [Program_Level=1.00] | 0<sup>a</sup> | . | . | . | . | . | . |
| [Time=3] *<br>[Program_Level=2.00] | -.155 | .2420 | -.630 | .319 | .413 | 1 | .521 |
| [Time=3] *<br>[Program_Level=1.00] | 0<sup>a</sup> | . | . | . | . | . | . |
| [Time=2] *<br>[Program_Level=2.00] | -.435 | .1996 | -.826 | -.044 | 4.748 | 1 | .029 |
| [Time=2] *<br>[Program_Level=1.00] | 0<sup>a</sup> | . | . | . | . | . | . |
| [Time=1] *<br>[Program_Level=2.00] | 0<sup>a</sup> | . | . | . | . | . | . |
| [Time=1] *<br>[Program_Level=1.00] | 0<sup>a</sup> | . | . | . | . | . | . |
| [Food_Insecurity=1] | .678 | .2900 | .109 | 1.246 | 5.462 | 1 | .019 |
| [Food_Insecurity=0] | 0<sup>a</sup> | . | . | . | . | . | . |
| (Scale) | 1 |  |  |  |  |  |  |

Dependent Variable: Q13\_PHQ-4 Score
Model: (Intercept), Time, Program\_Level, Time \* Program\_Level, Food\_Insecurity
a. Set to zero because this parameter is redundant.

Most clients identified as male for their gender identify representing 68.8% or 106 clients, while 26.0% of clients identified as female. Transgender female (4.5%) and other/non-conforming (0.6%) clients represented a smaller proportion of the client sample. 65.6% of clients identified as heterosexual regarding their sexual orientation representing 101 clients with the second largest group containing 32 clients representing 20.8%. Smaller proportions of sexual orientation include bisexual, other/non-conforming, pansexual, something else, chose not to respond, or had an unknown sexual orientation.

At baseline, the majority of clients were experiencing homelessness. Specifically, 140 clients or 90.9% of Harlem United clients were homeless at baseline and coded as “1”, while 14 clients, or 9.1% were not homeless at baseline and coded as “0.”

In this model, baseline is considered the reference category [Time = 1] and has been set to zero. There were 428 client housing records with 267 observations, or 62.4% classified as stable housing while 161 observations, or 37.6% were classified as unstable housing. The data was measured across three timepoints: baseline, year 1, and year 2. Baseline and year 1 both consisted of 154 observations representing 36.0% of the total observations. Year 2 consisted of 120 observations, representing 28.0% of observations. Year 2 contained missing housing data due to some clients having only one follow-up during Year 1.

A binomial logit GEE model was performed because the variable stable housing was coded as a binary variable. This approach was appropriate because it supports longitudinal data by accounting for repeated observations from the same client across time. The GEE model analyzed whether stable housing changed over time and was found to be statistically significant, Wald X^2^ (2) = 156.073, p < 0.001. The positive (B) values indicate that clients were more likely to have stable housing at Year 1 and Year 2 compared to the reference category at baseline. In Year 1, clients were shown to have 94.9% greater odds of stable housing compared to baseline with OR = 93.548, 95% Cl [45.176, 193.716], p < 0.001. In Year 2, clients had an increase of 94.9% greater odds of stable housing compared to baseline, OR = 94.875, 95% Cl [43.204, 208.345], p < 0.001. Findings suggest substantial improvement in housing status from baseline with Harlem United clients being provided with either permanent or temporary supportive housing.

Generalized estimating equations with a binomial distribution and logit link were used to assess changes in medication adherence over time and the effect of number of programs. A binomial logit GEE model was chosen because medication adherence was coded as a binary variable modeling the odds of adherence rather than medication adherence being a continuous number. This approach was appropriate because it supports longitudinal data by accounting for repeated observations from the same client across time. This model evaluated for effects of time, program level, and their interaction (time x program level) with the odds of adherence.

This model contained 233 total observations. There was no statistically significant change over time, Wald X^2^(2) = 0.005, p = 0.998. There was no statistically significant difference between medication adherence and program engagement, Wald X^2^ (1) = 2.428, p = 0.119. The interaction between time and program level was also not statistically significant, p = 0.469. There is no evidence that medication adherence changes over time based on program level. Medication adherence remained stable from baseline to year 2 with participation in programs not being associated with higher or lower medication adherence. When compared to baseline, there was no significant difference at Year 2, OR = 1.250, 95% Cl: 0.598-2.614, p = 0.553 or Year 1, OR = 0.926, 95% Cl: 0.590-1.454, p = 0.738. Clients involved in 3-5 programs compared to clients in 1-2 programs had lower odds of adherence, but were not statistically significant, OR = 0.458, 95% Cl: 0.117-1.799, p = 0.263.

A GEE model was used with a Poisson log distribution to evaluate PHQ4 scores for effects of time, program level, and their interaction (time x program level) and food insecurity. This approach was appropriate because the dependent variable is ordinal and supports longitudinal data by accounting for repeated observations from the same client across time. In this model, baseline is considered the reference category [Time = 1] and has been set to zero with data being measured across three timepoints: Baseline, Year 1, and Year 2.

In this model, 404 observations were observed. There was no statistically significant effect of time, Wald X^2^ (2) = 4.342, p = 0.114, suggesting PHQ4 scores did not significantly change over time. Program level was significantly associated with PHQ4 scores, Wald X^2^ (1) = 7.902, p = 0.005. Food insecurity was also associated with PHQ4 scores, Wald X^2^ (1) = 5.462, p = 0.019.

### Key Findings

The longitudinal data analysis among Harlem United clients showed several core outcomes that remained stable over time. Medication adherence and viral suppression remained stable across all time points with no statistically significant change over time. Stable housing improved from Baseline compared to Year 1 and Year 2 due to homelessness being inclusion criteria for both permanent and temporary supportive housing.^5^ These findings suggest that the Harlem United’s multiservice model supports the maintenance of key health and stability outcomes instead of large measurable improvements over time.

Harlem United provided several resources for both behavioral and mental health clients.^5^ According to the GEE model, PHQ4 scores did not significantly change over time, however, program level and food insecurity were significantly associated with PHQ4 scores. Clients involved with Harlem United multiservice program with greater behavioral health and social needs are more likely to be involved in multiple services. Food insecurity is another important factor that can impact anxiety and depression symptoms with food insecure clients having higher PHQ4 scores. This interpretation is supported by literature in the integration of HIV and mental health services.^5,6,8,14^ Integrated services have the potential to improve patient outcomes among people living with HIV with emphasis placed on implementation and service capacity.^14,17,18^ Findings from the longitudinal data analysis are supportive of existing literature showing that people living with HIV experience needs stemming from clinical, behavioral health, and social needs.^14,17,18^ Prior research has demonstrated that people living with HIV experience depressive symptoms related to HIV-related stigma and social support.^2,5–8^ Findings highlight the need for the continued push to incorporate interventions aimed at improving social support and resilience in mental health programs.^2,5–8^ Mental health outcomes are closely tied to both social and structural conditions with the noted association between food insecurity and PHQ4 scores in the GEE model. Food insecurity can be interpreted as a marker of chronic stress, economic instability, and a reduced capacity to prioritize one’s own health.^1–2,13–15,17,18^ Therefore, PHQ4 scores should be interpreted as a reflection of unmet social needs.^1–2^ Lastly, literature further analyzes how the different dimensions of social support are conducive to an effective multiservice model emphasizing the need for emotional, informational, and social support of people living with HIV.^1–2,13–15,17,18^

## Strengths

One major strength in the longitudinal analysis is it allows for the examination across Baseline, Year 1, and Year 2. This is an important strength because it provides the opportunity to evaluate the success of the program reflected through stability over time for clients with complex medical and social needs. Additionally, the use of the GEE model strengthens the analysis to account for within-client correlations rather than treating each observation independently.

## Limitations

Many of the outcomes included such as medication adherence and viral suppression were already high at baseline limiting room for improvement. Additionally, the GEE analysis cannot determine causality among clients with higher PHQ4 scores, even though it is suggested that clients with increased needs of behavioral health services were involved in more programs.

The study also did not have equal sample sizes with from Baseline, Year 1, and Year 2 due to some clients not continuing with the program after Year 1. Some GEE models had smaller observations which affected statistical significance and did not include for all possible confounders such as age, gender, or substance use. Including these variables in future analysis could strengthen the observed associations.

### Public Health Implications

Findings suggest that the multiservice model is supportive of HIV-related outcomes with maintaining stability through medication adherence and viral suppression. From a public health standpoint, maintaining high viral suppression rates is the goal to strengthen HIV health outcomes to ultimately achieve undetectable status.^3,6,13–15^ Programs serving people with HIV should incorporate screenings for food insecurity and nutrition support to better aid clients in reducing PHQ4 or behavioral symptoms.^6,13–15^ Due to clients facing challenges with both HIV and mental health, support of an integrated and client-centered program can impact more people living with HIV.^3,6,13–15,17,18^

### Future Direction

Future analyses should consider the type of services being offered and if outcomes were improved or maintained. Rather than grouping clients together, future direction could examine, food insecurity, harm reduction services, and behavioral health services more closely to determine association with PHQ4 score and viral suppression. Covariates would also be helpful to better understand client outcomes based on demographic characteristics demonstrating which clients benefited the most from multiservice engagement. Finally, qualitative reviews with staff could provide a deeper understanding into the client experience and how their needs were met.

## Conclusion

This study suggests that clients were transitioned into a stable housing from baseline through permanent or temporary housing. Viral suppression and Medication adherence maintained high levels throughout the duration of the program and did not represent statistical significance. The most meaningful findings were the impact of program level (number of programs) and food insecurity on PHQ4 scores. These findings demonstrate how clients who experienced food insecurity are more likely to experience depressive symptoms. Additionally, clients with higher PHQ4 scores were involved in more programs offered by Harlem United. Public health programs that serve people living with HIV should continue to strengthen integrated care models to further improve health outcomes related to HIV care.

## Data Availability

All data produced in the present work are contained in the manuscript

